# Perceptions of Complementary, Alternative, and Integrative Medicine Among Neurology Researchers and Clinicians: A Large-Scale, International Cross-Sectional Survey

**DOI:** 10.1101/2024.02.14.24302844

**Authors:** Jeremy Y. Ng, Stephanie Y. Li, Holger Cramer

## Abstract

**Background:** While many patients with neurological disorders and conditions use complementary, alternative, and integrative medicine (CAIM), little is known about the use and perceptions of CAIM among neurology researchers and clinicians. With the increasing popularity of CAIM, our objective was to assess practices, perceptions, and attitudes towards CAIM among neurology researchers and clinicians.

**Methods:** We conducted an anonymous online survey of authors who had published articles in neurology journals indexed in MEDLINE. We emailed potential participants our cross-sectional electronic survey after extracting their email addresses from one of their publications in our sample of journals. Basic descriptive statistics were drawn from quantitative data, and thematic content analysis was used to analyse qualitative data from any open-ended questions.

**Results:** The survey was completed by 783 neurology researchers and/or clinicians (1.5% response rate, 83.9% completion rate). Overall, respondents perceived CAIM to be promising in preventing, treating, and/or managing neurological diseases. Mind-body therapies received the most positive responses, indicated by over half of respondents cumulatively agreeing that they are promising (n=368, 59.0%) and safe (n=280, 50.3%). Whole medical systems and biofield therapy were less favourable. Most neurology clinicians reported a lack of formal (n=211, 70.3%) and supplementary training (n=158, 52.5%) on CAIM. Nearly half of clinicians did not feel comfortable counselling patients about CAIM (n = 121, 44.5%), and over half did not feel comfortable recommending it (n=161, 59.3%). A lack of scientific evidence for CAIM’s safety and efficacy was reported as the greatest challenge to CAIM (n=515, 92.5%). The majority of respondents believed there is value to conducting research on this topic (n=461, 82.0%) and supported increasing allocation of research funding towards CAIM (n=241, 58.9%).

**Conclusions:** Although many participants found CAIM to be promising to the field of neurology, the vast majority did not feel open to integrating CAIM into mainstream medical practices on account of a perceived lack of scientific evidence for its safety and efficacy. Future studies can use our findings to improve educational resources on CAIM within neurology, as well as examine what effects a tailored CAIM education has on the perceptions of neurology researchers and clinicians.

## Introduction

Neurology is a vital field of medicine focused on the diagnosis and treatment of disorders specific to the nervous system^1,2^. Beyond its core structural components—including the brain, spinal cord, and peripheral nerves—the nervous system is central to controlling and coordinating all bodily functions, from the consolidation of memories to the beating of the heart^3^. Given its cascade of functional influence over both physical and psychological systems^3^, neurological disorders collectively represent the leading cause of disability and the second leading cause of death, impacting millions of individuals worldwide^4^. Globally, 10 million deaths and 349 million disability adjusted life years were estimated to arise from neurological disorders in 2019^5^.

While conventional medical treatments are available, chronic neurological conditions typically prove challenging to treat^6^. Although many neurological conditions are common^6,7^—including headache and migraine^8-12^, depression^13-16^, chronic pain^17^, multiple sclerosis^18-20^, and Parkinson’s disease^21^—conventional, evidence-based medicine such as pharmacotherapy often only achieves symptom relief, or is associated with adverse side effects^6-18,20-25^. Several studies report that individuals suffering from chronic conditions choose to seek complementary, alternative, and integrative medicine (CAIM)^24,27^ as a means of actively reducing specific symptoms associated with their condition, as well as improving general health and wellness^6,9,10,11,14,18,19,22,23,25,27,28^.

According to the National Center for Complementary and Integrative Health (NCCIH), “complementary medicine” encompasses non-mainstream approaches used in conjunction with conventional medicine, while “alternative medicine” refers to non-mainstream approaches used in place of conventional medicine. “Integrative medicine” encompasses a holistic combination of conventional and complementary medicines that emphasizes caring for all aspects of a patient’s health; this includes their biological, behavioural, social, and environmental well-being^25,27^. For the purpose of this study, we will collectively refer to this group of diverse therapies as CAIM.

The prevalence of CAIM use among patients with chronic neurological conditions is consistently described in literature as widespread^6-28^. It was previously reported that adults with common neurological conditions use CAIM more frequently than those without (44.1% vs 32.6%)^10^. The most common CAIM therapies used by patients with neurological conditions include biologically based practices, such as herbal and dietary supplements; body-based practices, including chiropractic therapy and massage; mind-body therapies, including meditation, yoga, and biofeedback; and whole medical systems such as traditional Chinese medicine^6-29^. Most patients with chronic conditions use CAIM therapies concurrently with conventional medicine^9,15^, and self-report some perceived benefit and symptom relief^7,9,10,11,19,23,25,28,31,32^. Despite the growing interest and patient demand for CAIM, there is a lack of high-quality literature supporting the safety and efficacy of using CAIM alongside conventional medicine^6-16,18-23,25-30,32-42^. For example, a large number of trials that compare Ginkgo (*Ginkgo biloba*) to placebos have reported inconsistent outcomes and conclusions on its efficacy for patients with dementia and/or cognitive impairment^33^. Numerous publications have also documented significant drug-drug interactions between Ginkgo and conventional medicine^35,36^. Correspondingly, investigation into potential interactions between CAIM and conventional medicine is scant across all neurological conditions^13^. This presents an issue of patient safety, exacerbated by data that suggests that over 50% of patients with neurological conditions do not disclose their CAIM usage with their health care provider^6^. In consideration of the prevalence of CAIM usage among patients with neurological conditions, its discrepancy with the prevalence of evidence-based CAIM therapies^10^, and the lack of clinician-patient communication surrounding CAIM, there is a clear issue regarding treatment safety and the overall outcome of patient care^6,8,10,13-15,17,19,34^. Although clinicians’ acceptance of CAIM has notably increased over the last two decades, their knowledge of and experience with many CAIM treatments largely remains unchanged. This is likely due to the lack of reliable evidence for many CAIM therapies pertaining to neurological conditions that prevents it from being included in most clinical practice guidelines^12,16,18,34,37-40^. The majority of physicians across all medical specialities have reported feeling uncomfortable recommending or discussing CAIM with their patients as a result^16,37,38^.

To our knowledge, no studies have investigated the perceptions of CAIM among neurology researchers and clinicians. These insights may aid in identifying potential barriers to clinician-patient communication surrounding CAIM within a neurological context, as well as emphasize a potential disconnect between neurology clinicians and their patients^34,37-42^. Our findings may identify underlying reasons for the apparent bias against CAIM therapies among neurology researchers and clinicians^37-42^, and consequently, warrant the potential need for CAIM to be prioritized in clinical, research, and educational initiatives^41,42^. In doing so, the needs and interests of patients with neurological conditions may be better reflected by the healthcare system, and neurology clinicians can make more informed, evidence-based decisions when integrating CAIM with conventional medicine^12,16,18,42^.

In this study we sought to investigate neurology researchers’ and clinicians’ perceptions of CAIM and CAIM categories, specifically, mind-body therapies, biologically based practices, manipulative and body-based practices, biofield therapy, and whole medical systems. These findings may offer a comprehensive understanding of the current knowledge and use of CAIM in conventional treatment settings for patients with neurological conditions^12,16,18,37-42^.

## Methods

### Transparency Statement

Clearance from the University Hospital Tübingen Research Ethics Board was obtained prior to starting this project (REB Number: 389/2023BO2). The study protocol was registered and made available on Open Science Framework (OSF)^43^ prior to recruiting participants. The study materials and raw data were shared on OSF^44^.

### Study Design

An anonymous cross-sectional survey was conducted online with a complete sample of authors who have published articles in neurology journals indexed in MEDLINE^45^. Based on the nature of the search, the majority of these authors were inferred to be neurology researchers and/or clinicians.

### Sampling Framework

For the purpose of this study, a complete sample of corresponding authors who have published articles in neurology journals within the last 5 years were considered potential participants. First, NLM IDs of all publications under the broad subject term ‘Neurology[st]’ were extracted (.txt) from the NLM catalog and used to develop a search strategy; the complete list of neurology journals can be found at the following link: https://journal-reports.nlm.nih.gov/broad-subjects/^46^. A copy of the search strategy is available on OSF (see: https://osf.io/hmkqs). Next, the search strategy was executed on OVID MEDLINE to yield a list of PMIDs that were exported in batches of 2000 (.csv). Each batch was inputted through a custom R script that accessed easyPubMed^47^ and outputted pertinent author data, including name, affiliated institution(s), and email address(es). A power analysis was not included on account of the use of a convenience sample with descriptive work and the lack of inferential testing.

### Participant Recruitment

Data cleaning of the email list output by our sampling framework was performed to mitigate the risk of repeated and unintended recruitment. All remaining authors were invited to participate in this study. Participants were contacted by email using SurveyMonkey^48^. The invitation included an authorized recruitment script outlining this study, its objectives, and a link to the survey. The survey link initially redirected participants to an implied consent form that was required to be acknowledged prior to participation. Upon starting the survey, respondents were met with a screening question that determined their eligibility to participate; researchers within the field of neurology (i.e., neurology was one of their fields of expertise) and/or clinicians within the field of neurology (i.e., healthcare providers who specialized in/had a practice focused on neurology) were considered eligible. During the survey, participants had the option to skip any questions that they did not wish to answer. To encourage participation, weekly reminder emails were dispatched for three weeks following initial contact, and potential participants were given a total of 8 weeks to choose to participate. Responses were collected from August 8, 2023, to October 3, 2023. There was no financial compensation and no requirement to participate in this study.

### Survey Design

SurveyMonkey^48^ was used to build and administer the survey. The complete survey is available on OSF (see: https://osf.io/3jz9c). The survey contained 33 questions and was displayed across 8 pages (screens). Respondents were first met with a screening question that determined their eligibility to participate; ineligible users were disqualified from continuing the survey and were redirected to an exit screen. Eligible users continued to answer a set of general demographic questions including gender, clinical/research role(s), career stage, and region of residence. All remaining questions focused on capturing respondents’ perspectives on CAIM using multiple-choice and open-ended questions. Two independent CAIM researchers pilot tested the survey prior to distribution. Their feedback was considered and integrated in the final survey version.

### Data Management and Analysis

This study had no formal hypotheses. An analysis of quantitative data yielded basic descriptive statistics. With respect to qualitative data, thematic content analysis was used to identify repeated ideas from the open-ended questions. The final set of codes was categorized and grouped into themes by our research team^49^. For reporting purposes, the finalized codes were used to categorically organize the data into tabular format. The Checklist for Reporting Results of Internet E-Surveys (CHERRIES) was used to inform the reporting of this survey^50^.

## Results

### Search Results

After the search strategy was executed on OVID MEDLINE for records published between October 1, 2020, and May 1, 2023, 123 874 articles were returned. From these articles, 123 389 PMIDs were obtained. A total of 54 875 email addresses were extracted using easyPubMed, of which 3796 duplicates were identified and removed from the raw dataset. The final, cleaned email list contained 44 987 unique email addresses and was used to recruit participants. The raw (deidentified) survey data is available here: https://osf.io/mtcea.

### Study Flow and Participant Demographics

Our survey yielded 783 participants, with a response rate of 1.7% (total responses ÷ opened and unopened invitations) and a completion rate of 83.9% (n=657). Incomplete responses were defined as responses with no questions answered following the initial screening questions. The raw response rate we have presented is an underestimation as we cannot determine how many of the 44 987 authors who were emailed presently identify as a neurology researcher and/or clinician. An opened rate of 42.2% (n=18,984) and an unopened rate of 58.1% (n=26,149) were calculated. Moreover, an email bounce back rate of 11.9% (n=6,090) was calculated. The response rate of opened invitations was 4.1%. The survey took 9 minutes and 10 seconds to complete on average. All questions were optional, hence we have provided the individual response rate of each question in parentheses.

In total, 761 participants self-identified as one of the following within the field of neurology: researcher only (n = 339, 44.5%), researcher and clinician (n = 297, 39%), or clinician only (n = 34, 4.5%). Respondents were primarily located in Europe (n = 297, 45.1%) or the Americas (n = 230, 35.0%), and the majority identified as a senior career researcher/clinician (n = 387, 59.0%) holding the position of faculty member/principal investigator (n = 352, 53.5%). A sizeable proportion of respondents also identified as a clinician (n = 222, 33.7%) or scientist in academia (n = 205, 31.2%). The majority of researchers were involved in clinical research (n = 406, 67.9%). Three in five neurology researchers (n = 360, 60.7%) had never conducted any form of CAIM research. Complete participant demographics are described in **Table 1**.

**Table 1.** Characteristics of Survey Participants.

|  |  |
| --- | --- |
| **Sex (n = 657)** |  |
| Male | 377 (57.4%) |
| Female | 267 (40.6%) |
| Other | 13 (2%) |
| **Age (n = 658)** |  |
| 18-24 | 2 (0.3%) |
| 25-34 | 88 (13.4%) |
| 35-44 | 191 (29%) |
| 45-54 | 181 (27.5%) |
| 55-64 | 121 (18.4%) |
| >65 | 69 (10.5%) |
| Prefer not to say | 6 (0.9%) |
| **Visible Minority (n = 652)** |  |
| Yes | 98 (15%) |
| No | 525 (80.5%) |
| Prefer not to say | 29 (4.5%) |
| **Location (n = 658)** |  |
| Africa | 5 (0.8%) |
| Americas | 230 (35%) |
| Eastern Mediterranean | 27 (4.1%) |
| Europe | 297 (45.2%) |
| South-East Asia | 43 (6.5%) |
| Western Pacific | 43 (6.5%) |
| Prefer not to say | 13 (2%) |
| **Current Position (n = 658)** |  |
| Clinical Student | 6 (0.9%) |
| Clinician | 222 (33.7%) |
| Graduate Student | 21 (3.2%) |
| Postdoctoral fellow | 70 (10.6%) |
| Faculty member/principal investigator | 352 (53.5%) |
| Research support staff | 26 (4%) |
| Scientist in academia | 205 (31.2%) |
| Scientist in industry | 12 (1.8%) |
| Scientist in third sector | 9 (1.2%) |
| Government scientist | 16 (2.4%) |
| Other | 14 (2.1%) |
| **Career Stage (n = 656)** |  |
| Graduate student | 12 (1.8%) |
| Early career researcher (<5 years post education) | 113 (17.2%) |
| Mid-career research (5-10 years post education) | 144 (22%) |
| Senior researcher (>10 years post education) | 387 (59%) |
| **Primary Research Area (n = 598)** |  |
| Clinical research | 406 (67.9%) |
| Preclinical research (in vivo) | 191 (31.9%) |
| Preclinical research (in vitro) | 110 (18.4%) |
| Health systems research | 44 (7.4%) |
| Health services research | 58 (9.7%) |
| Methods research | 85 (14.2%) |
| Epidemiological research | 107 (17.9%) |
| Other | 33 (5.5%) |
| **Area of CAIM Research (n = 593)** |  |
| Mind-body therapies | 83 (14%) |
| Biologically based practices | 128 (21.6%) |
| Manipulative and body-based practices | 26 (4.4%) |
| Biofield therapy | 6 (1%) |
| Whole medical systems | 29 (4.9%) |
| Never conducted any CAIM research | 360 (60.7%) |
| Other | 37 (6.2%) |

### Perceptions about CAIM

Participants were asked to identify all CAIM categories they perceived to be the most promising in preventing, treating, and/or managing neurological diseases or conditions. The categories were: mind-body therapies, biologically based practices, manipulative and body-based practices, biofield therapy, and whole medical systems. Of the 624 respondents, over half expressed positive attitudes toward mind-body therapies (n = 368, 59.0%). Many respondents also perceived most biologically based practices (n = 292, 46.8%), manipulative and body-based practices (n = 123 19.7%), and whole medical systems (n = 119, 19.1%) to be promising. Eighty-three (13.8%) respondents stated that they did not perceive any CAIM categories to be promising.

We then asked to what extent respondents agree that each CAIM category is safe, using a 5-point Likert scale. Respondents were roughly evenly split between “agree” (n = 213, 37.7%) and “neither agree nor disagree” (n = 200, 35.4%) regarding CAIM therapies in general. Half of the respondents agreed that most mind-body therapies are safe (n = 280, 50.3%). However, the majority remained neutral to the remaining CAIM categories: biologically based practices (n = 229, 41.1%), manipulative and body-based practices (n = 199, 35.7%), biofield therapy (n = 247, 44.9%), and whole medical systems (n = 233, 42.1%) (**Figure 1**).

**Figure 1.**
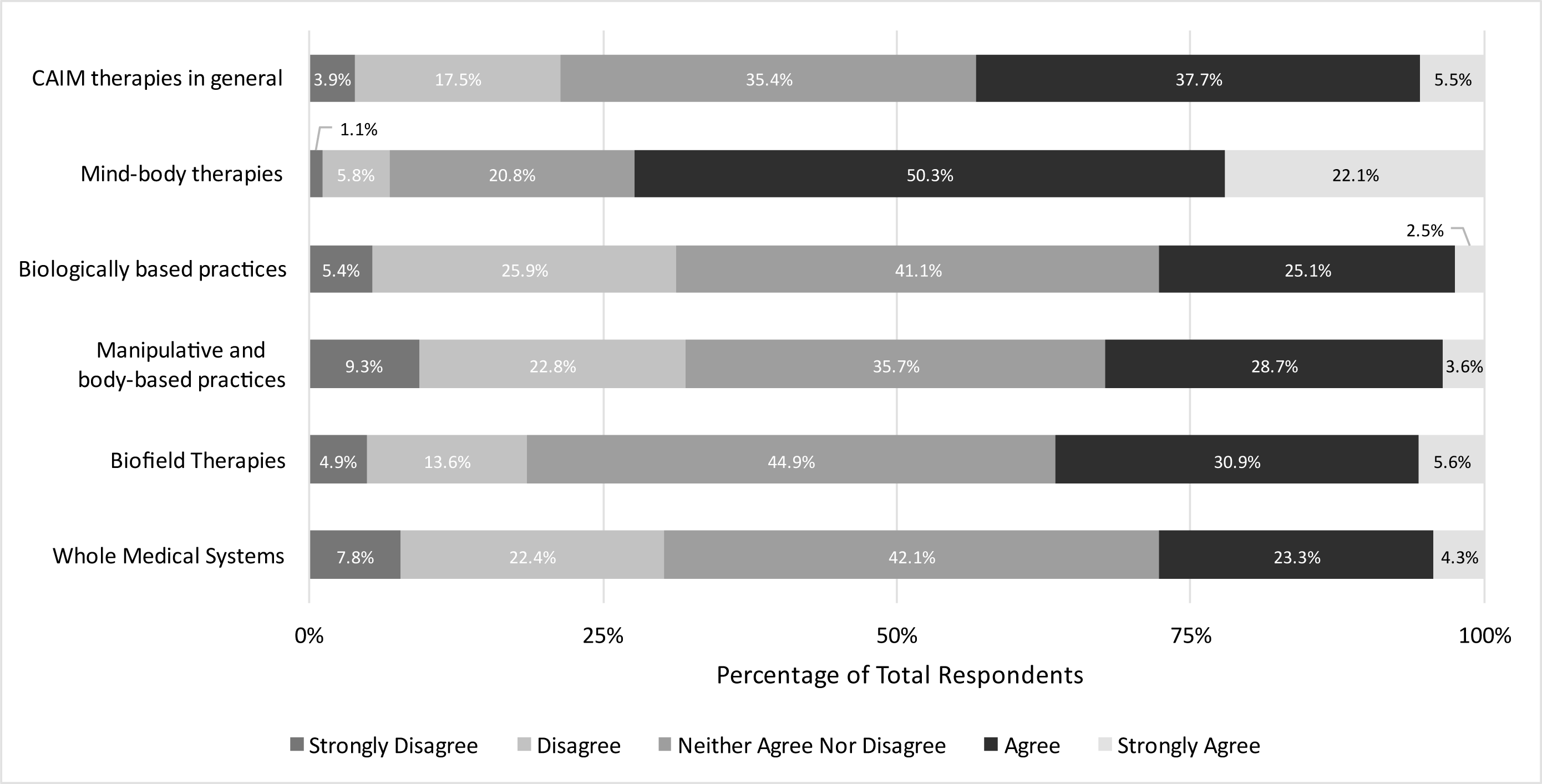
Participants’ Agreement Expressed Towards the Safety of Each CAIM Category.

In addition, we asked participants to what extent they agreed that each CAIM category is effective. Approximately two-fifths of respondents remained neutral across all categories, as well as CAIM in general. More than one third of respondents agreed or strongly agreed that most mind-body therapies are effective (n = 201, 36.2%). In contrast, higher levels of disagreement (either “disagree” or “strongly disagree”) were expressed towards CAIM therapies (n = 273, 48.5%), particularly biologically based practices (n = 221, 39.8%), manipulative and body-based practices (n = 180, 32.3%), biofield therapy (n = 285, 51.8%) and whole medical systems (n = 232, 42.0%) (**Figure 2**).

**Figure 2.**
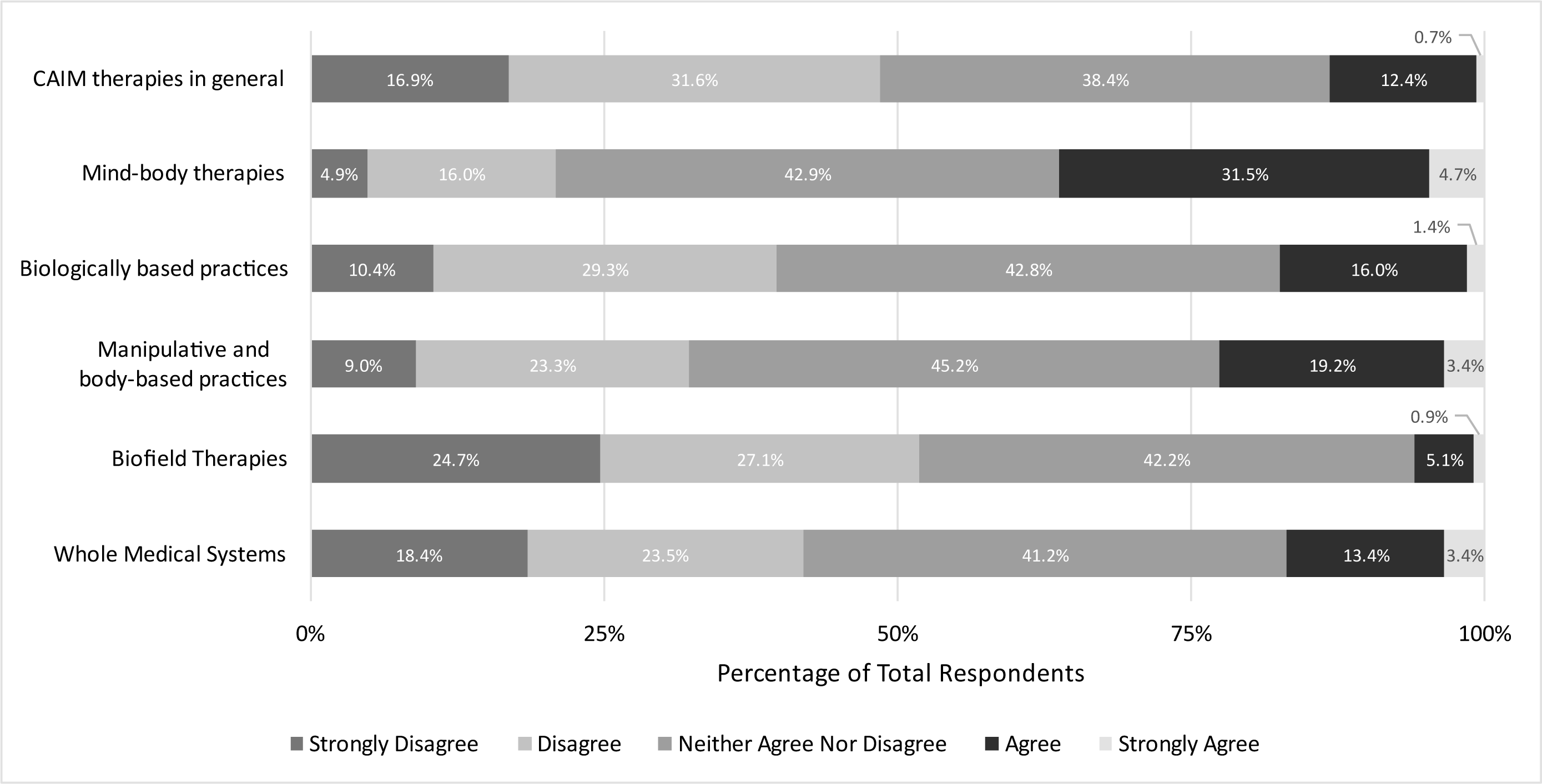
Participants’ Agreement Expressed Towards the Efficacy of Each CAIM Category.

The top benefits respondents associated with CAIM included: “focus on prevention and lifestyle changes” (n = 354, 64.5%), “expanded treatment options” (n = 345 63.2%), “holistic approach” (n = 323, 59.2%), and “patient empowerment” (n = 279, 51.1%). Additionally, nearly half of respondents believed CAIM holds “cultural and spiritual relevance” (n = 262, 48.0%), “increases patient satisfaction and well-being” (n = 262, 48.0%), and has the “potential to address chronic health conditions that conventional medicine has been unable to treat effectively” (n = 258, 47.3%) (**Figure 3**). Despite these benefits, 515 of 557 respondents (92.5%) expressed that the lack of scientific evidence for CAIM’s safety and efficacy is a challenge. In addition, “lack of standardization in product quality and dosing” (n = 515, 86.0%), “difficulty in distinguishing legitimate practices from scams or fraudulent claims” (n = 479, 76.7%), and “limited regulation and oversight” (n = 427, 71.6%) are all notable barriers associated with CAIM. Moreover, 42.7% (n = 238) of respondents perceived high costs and lack of insurance coverage to be a challenge (**Figure 4**), and were evenly split between favouring, disfavouring, and remaining neutral to the idea that insurance companies should cover the costs of most CAIM therapies.

**Figure 3.**
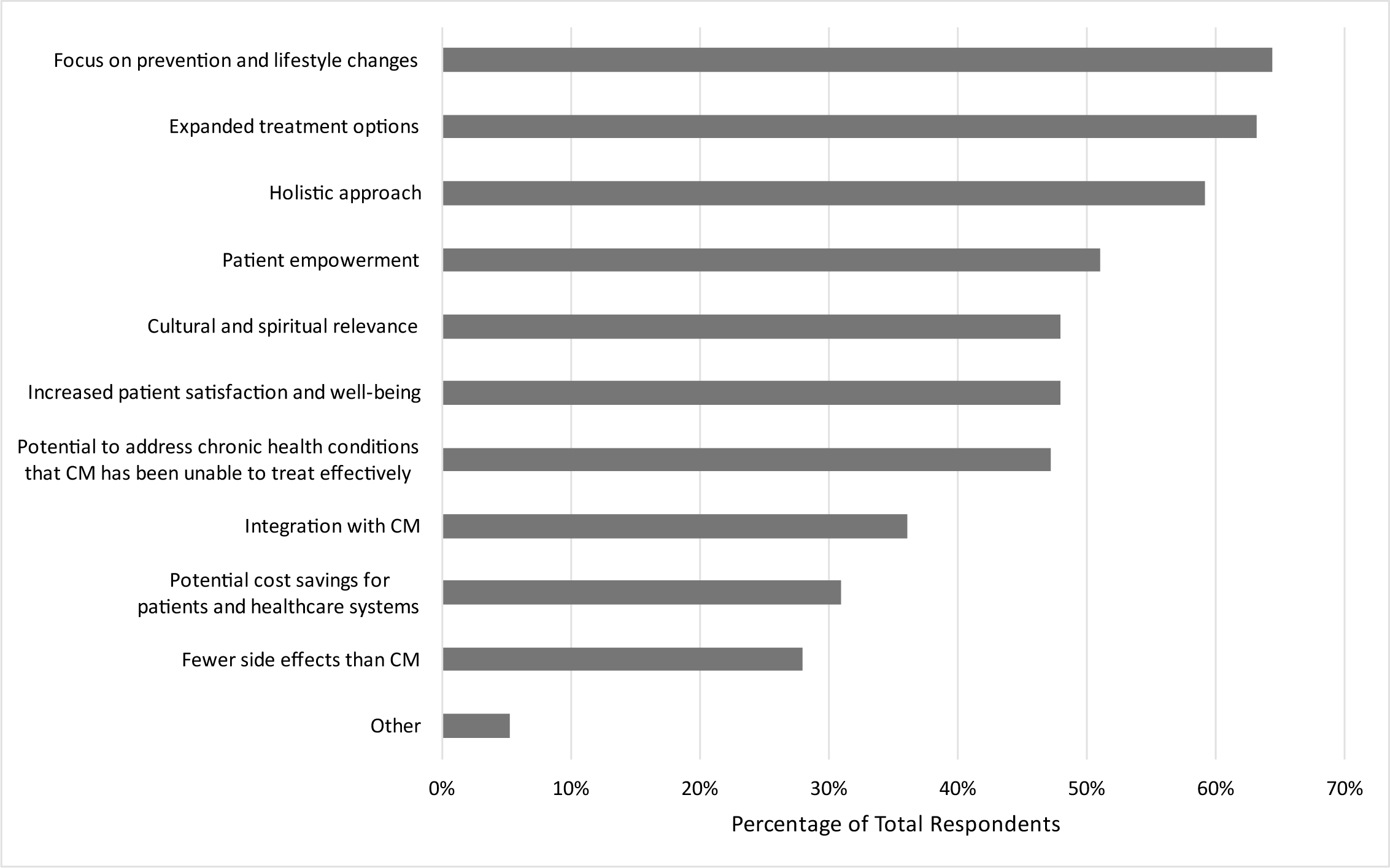
Benefits Participants Associated with CAIM.

**Figure 4.**
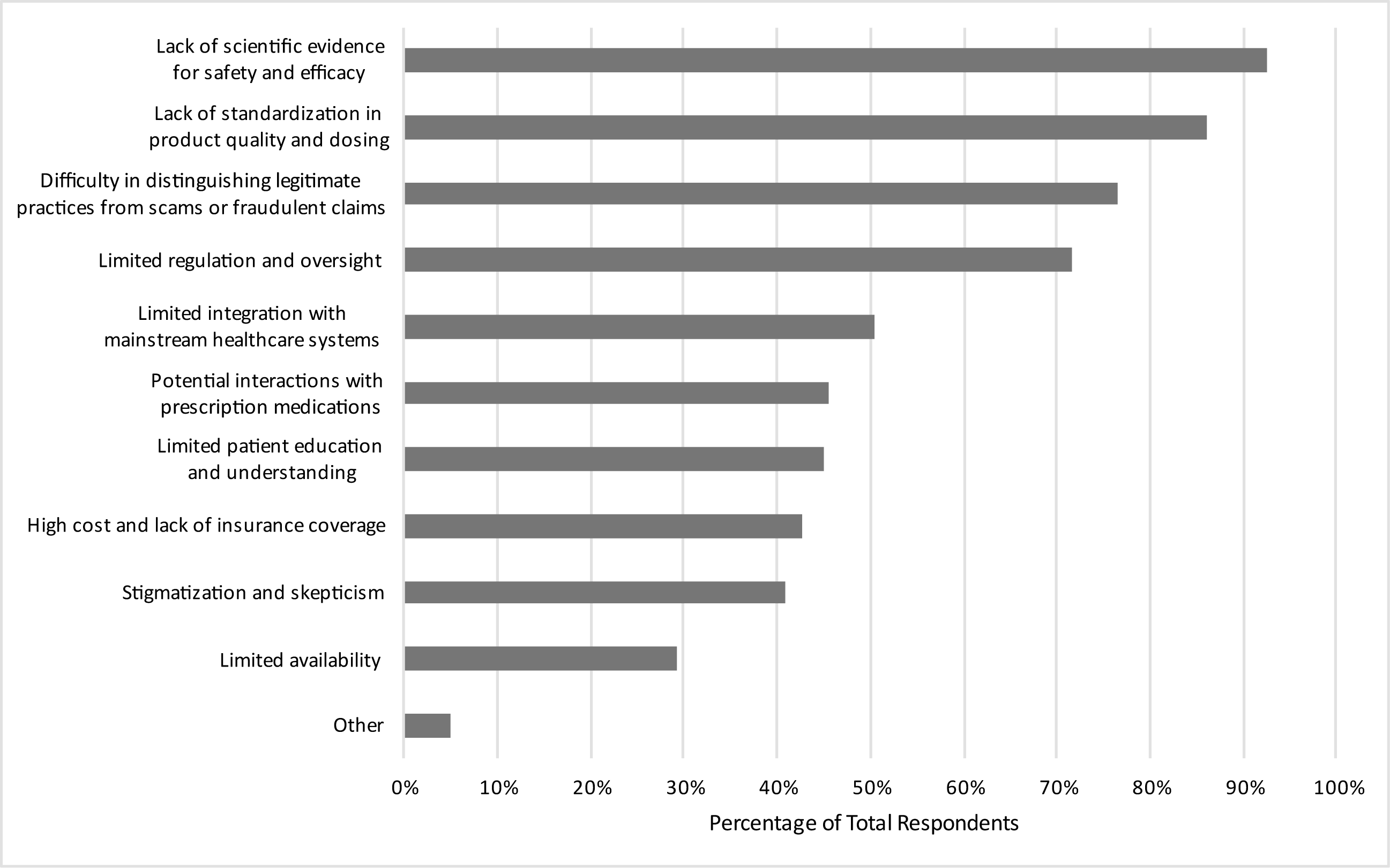
Challenges Participants Associated with CAIM.

### Clinical Education and Experiences with CAIM

Among neurology clinicians, 211 of 300 respondents (70.3%) never received formal training in any area of CAIM. Over half (n = 158, 52.5%) of respondents report having not received any supplemental training either. Of those who have received formal and/or supplemental training, mind-body therapies (n = 47, 15.7%; n = 96, 31.9%) and biologically based practices (n = 45, 15.0%; n = 84, 27.9%) were the most prevalently studied categories.

More than three-fifths of clinicians have experienced patient-initiated discussions concerning biologically based practices (n = 244, 80.26%), mind-body therapies (n = 225, 74.0%), manipulative and body-based practices (n = 196, 64.5%), and/or whole medical systems (n = 191, 62.8%). Additionally, one fifth of respondents reported engaging with patients about biofield therapy (n = 61, 20.1%). Although 93.4% of clinicians (n = 284) have had patients seek counselling or disclose using CAIM in the past, in last 12 months alone, the majority of clinicians (n = 171, 56.3%) only recall 0-20% of patients doing so. Of this majority, most responses fall in the 0-10% range (n = 95, 31.3%).

Furthermore, when asked to express the extent to which they are comfortable counselling patients about each CAIM category, mind-body therapies was the only intervention that the majority of respondents felt comfortable with (n = 140, 51.9%). Approximately two-fifths of clinicians did not feel comfortable counselling patients about CAIM (n = 121, 44.5%), more specifically, manipulative and body-based practices (n = 108, 40.3%), biofield therapy (n = 154, 57.57%), and whole medical systems (n = 136, 50.8%) (**Figure 5**).

**Figure 5.**
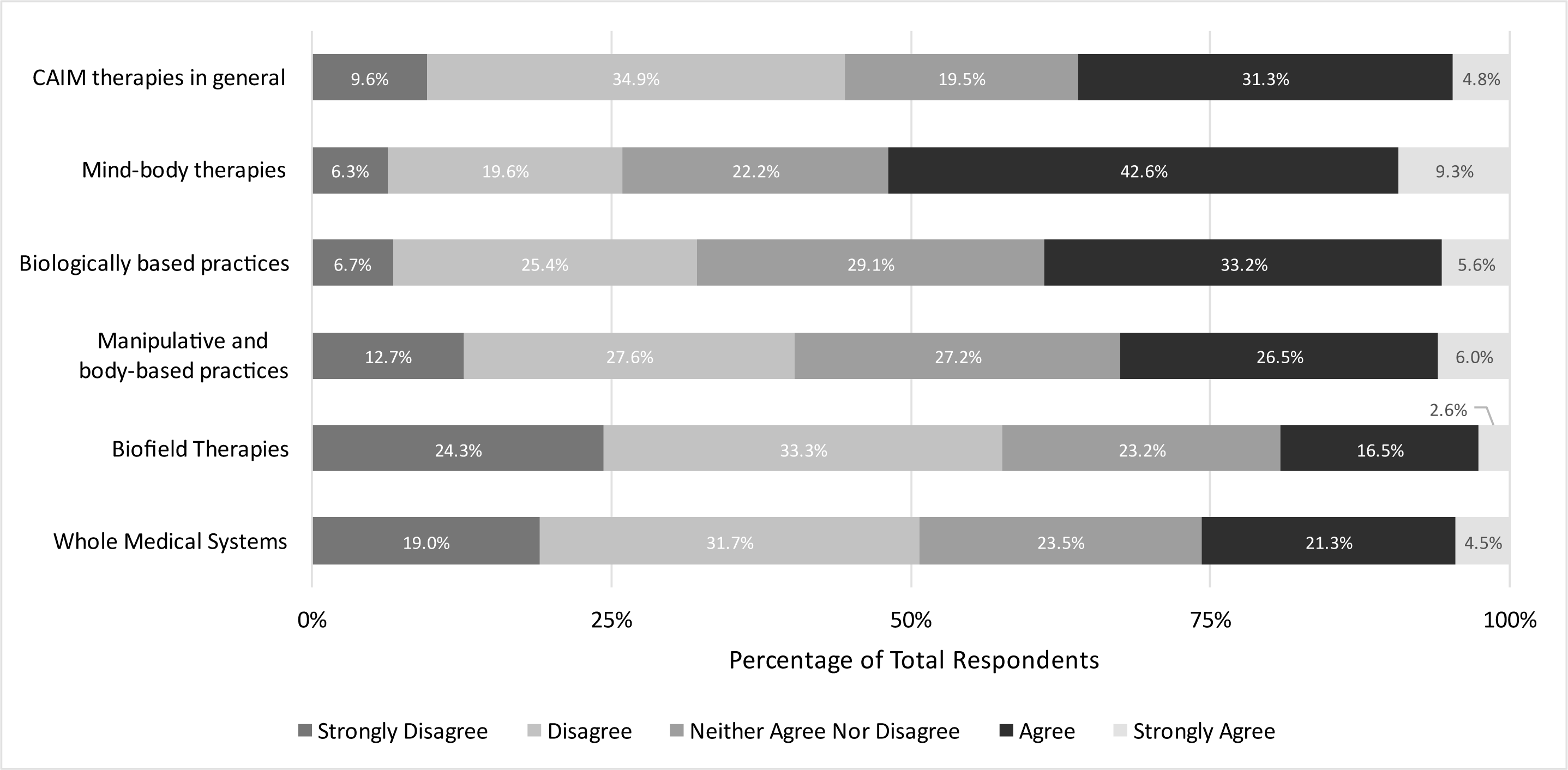
Clinicians’ Agreement Expressed Towards Feeling Comfortable Counselling Patients on Each CAIM Category.

Nearly 80% of clinicians report having practiced or recommended CAIM to their patients. Mind-body therapies (n = 179, 58.7%) and biologically based practices (n = 125, 41.0%) were the most recommended therapies. Interestingly, when clinicians were asked how comfortable they would feel recommending each CAIM category to patients, responses were more varied. Approximately 44% (n = 118) of clinicians would be comfortable recommending mind-body therapies to their patients. In contrast, over 50% of respondents cumulatively disfavoured recommending CAIM therapies in general (n = 161, 59.3%), particularly biologically based practices (n = 129, 48.3%), manipulative and body-based practices (n = 144, 53.7%), biofield therapies (n = 187, 69.8%), and whole medical systems (n = 159, 59.3%) (**Figure 6**).

**Figure 6.**
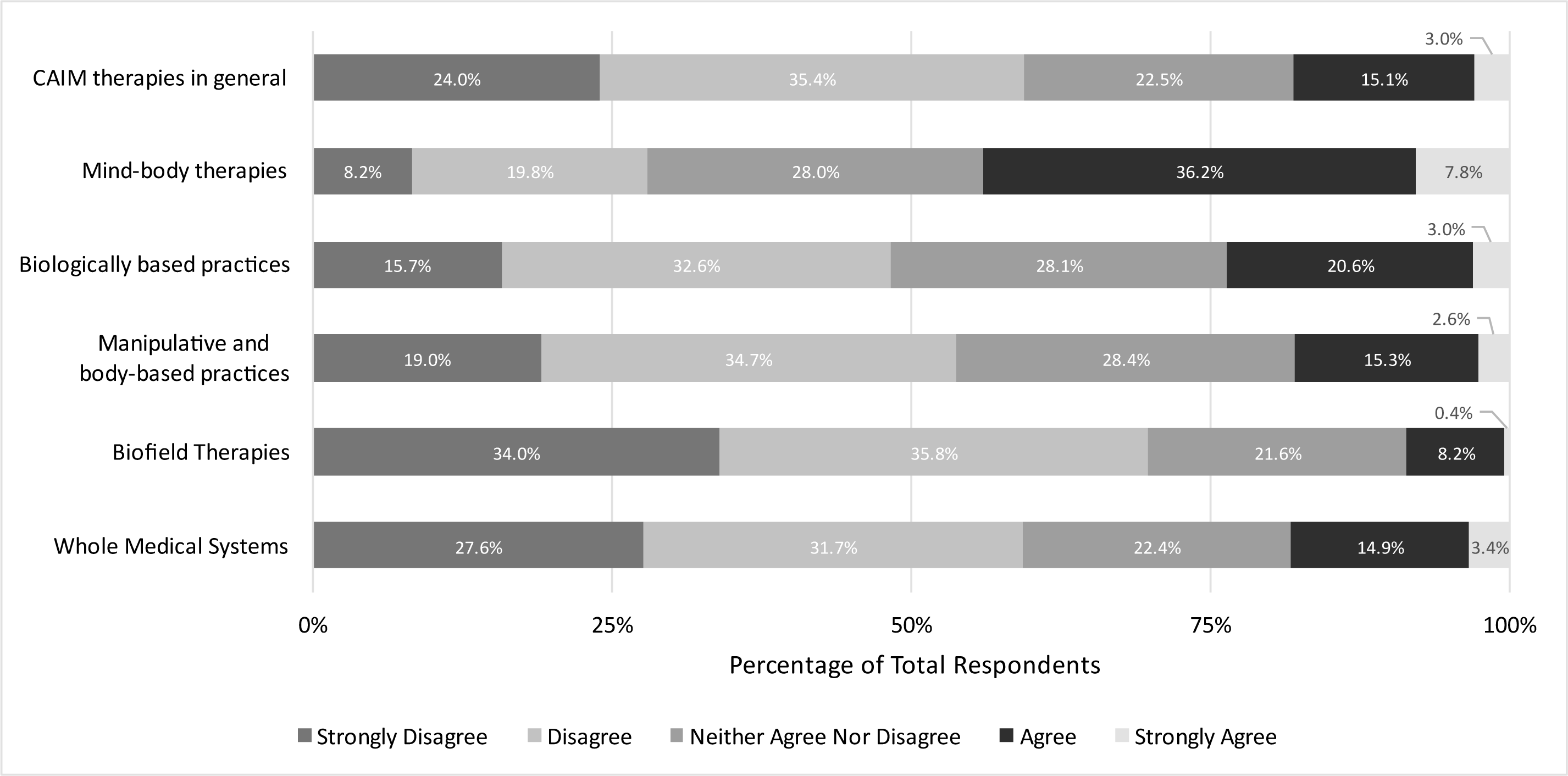
Clinicians’ Agreement Expressed Towards Feeling Comfortable Recommending Each CAIM Category to Patients.

### Attitudes towards CAIM Training and Practices

When asked whether each CAIM category should be integrated into mainstream medical practices, participants’ opinions were relatively varied. Between 30-40% of respondents remained neutral across all categories. Respondents were evenly split between disfavouring, favouring, and remaining neutral to biologically based practices and manipulative and body-based practices. Greater levels of disagreement were expressed towards CAIM as a whole (n = 157, 40.4%), particularly biofield therapies (n = 287, 52.1 %) and whole medical systems (n = 235, 42.5 %). Mind-body therapies was the most supported CAIM category regarding integration into mainstream medical practices (n = 222, 39.9%) (**Figure 7**).

**Figure 7.**
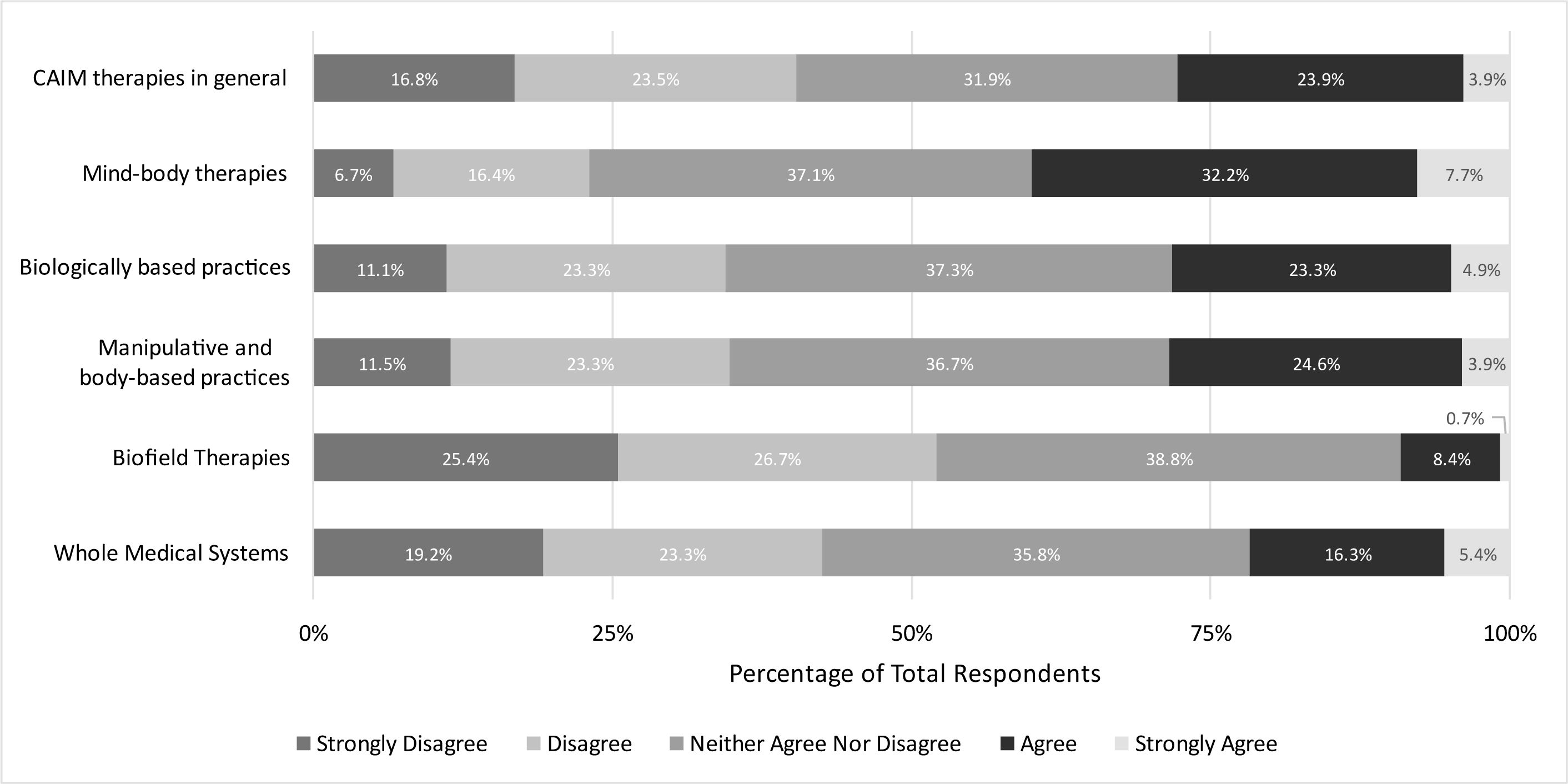
Participants’ Agreement Expressed Towards the Integration of Each CAIM Category Into Mainstream Medical Practices.

With respect to clinical training, more than half of respondents cumulatively agreed that clinicians should receive formal education on CAIM (n = 308, 54.7%), particularly mind-body therapies (n = 302, 54.2%) and biologically based practices (n = 313, 56.2%) (**Figure 8**). Similar sentiments were expressed towards these categories regarding supplementary education; approximately three in five respondents cumulatively agreed that clinicians should receive supplementary training on CAIM (n = 350, 62.1%), specifically, mind-body therapies (n = 344, 61.7%) and biologically based practices (n = 324, 58.2%) (**Figure 9**). Most respondents either agreed or remained neutral to formal clinical training on manipulative and body-based practices (n = 205, 36.8%; n = 164, 29.6%) and whole medical systems (n = 189, 33.9%; n = 172, 31.1%) (**Figure 8**). Interestingly, a greater proportion of respondents supported supplementary training for these categories (n = 260, 46.9%; n = 241, 43.6%) (**Figure 9**). Regarding biofield therapy, 43.0% of respondents did not think clinicians should receive formal education on this topic (n = 237) (**Figure 8)** However, responses were more evenly split when it came to supplementary education (**Figure 9**).

**Figure 8.**
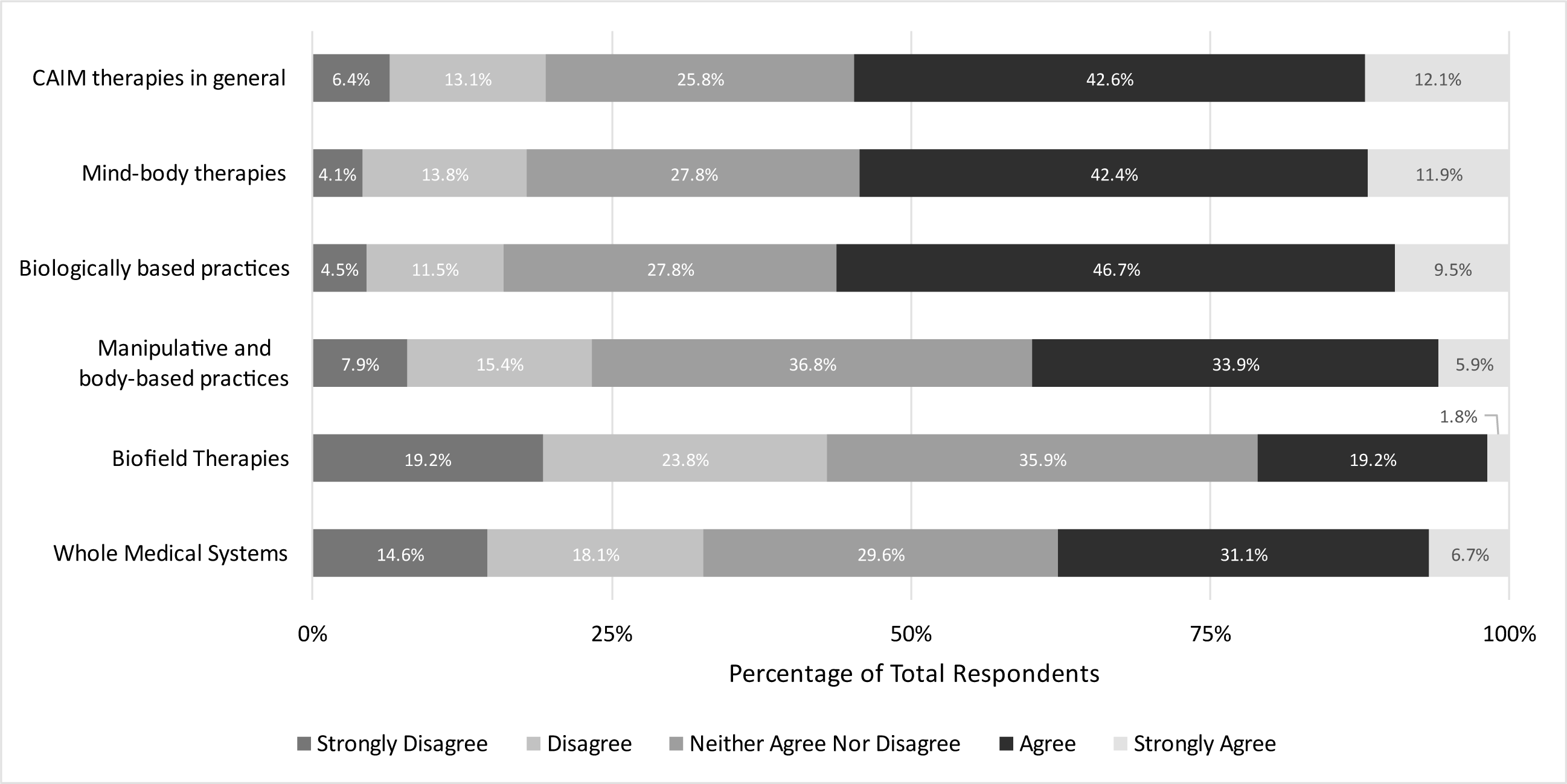
Participants’ Agreement Expressed Towards Clinicians Receiving Formal Education on Each CAIM Category.

**Figure 9.**
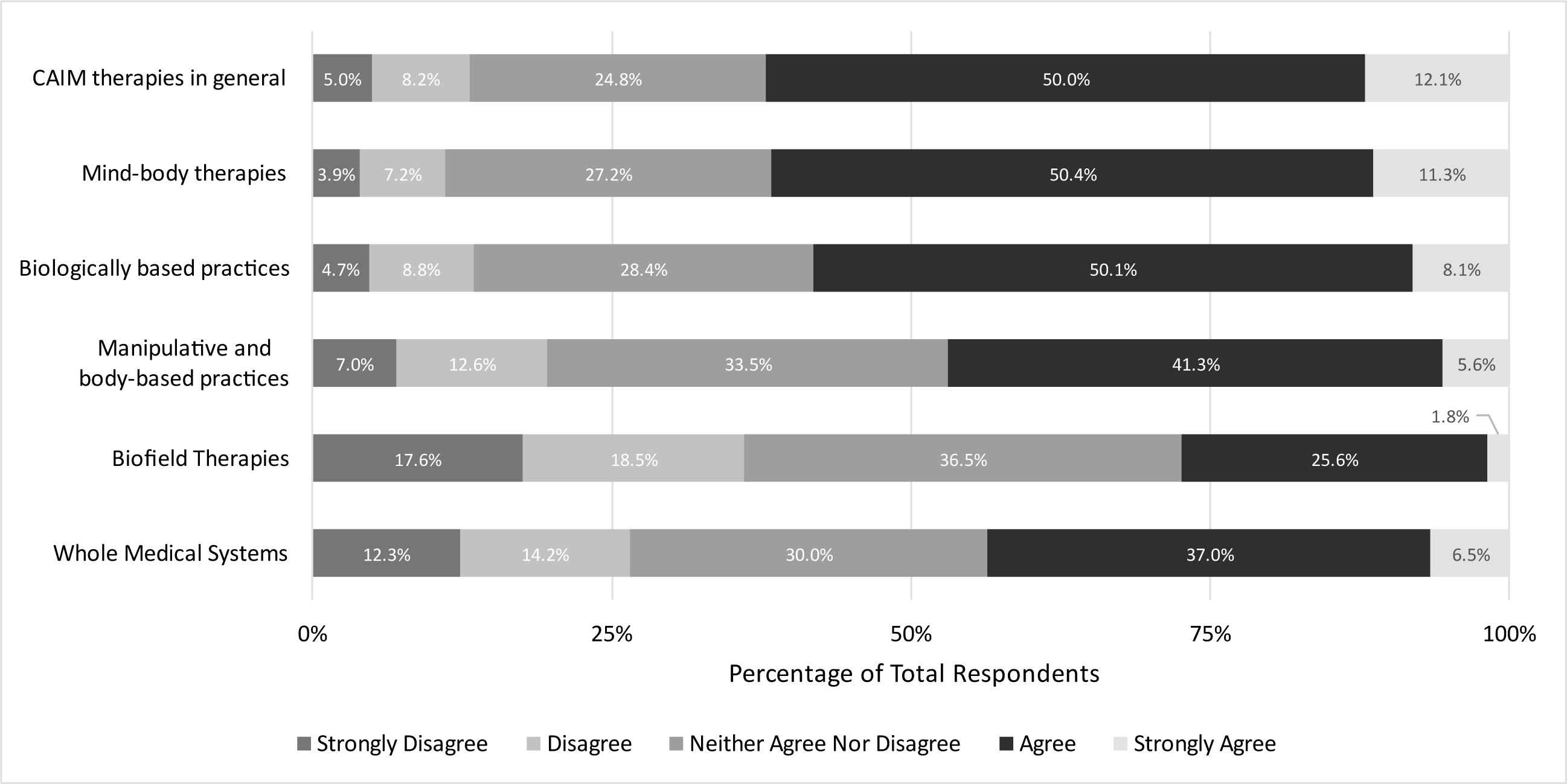
Participants’ Agreement Expressed Towards Clinicians Receiving Supplementary Education on Each CAIM Category.

### Attitudes towards CAIM Research

The vast majority of researchers and clinicians reported that they would seek out academic literature (n = 578, 92.3%) to learn more about CAIM. Other notable sources included conference presentations or workshops (n = 265, 42.3%), colleagues (n = 216, 34.5%), and health or information pages on the internet (n = 201, 32.1%). In addition, over three-quarters of respondents agreed or strongly agreed that there is value to conducting research on CAIM (n = 461, 82.0%), particularly mind-body therapies (n = 452, 80.9%) and biologically based practices (n = 427, 76.7%). More than 60% of respondents felt similarly about manipulative and body-based practices (n = 356, 64.0%) and whole medical systems (n = 335, 60.6%).

Respondents were evenly divided between disfavouring, favouring, and remaining neutral to research on biofield therapies (**Figure 10**). When asked to what extent they agreed with allocating more research funding towards CAIM, many respondents agreed (n = 121, 37.6%) or strongly agreed (n = 120, 21.3%). Across specific CAIM categories, over half of respondents favoured mind-body therapies (n = 332, 59.5%) and biologically based practices (n = 317, 57.0%). Approximately 45% of respondents agreed or strongly agreed that manipulative and body-based practices (n = 252, 45.5%) and whole medical systems should receive more research funding (n = 250, 45.1%). Respondents were evenly split between disfavouring, favouring, and remaining neutral to allocating funding towards biofield therapy.

**Figure 10.**
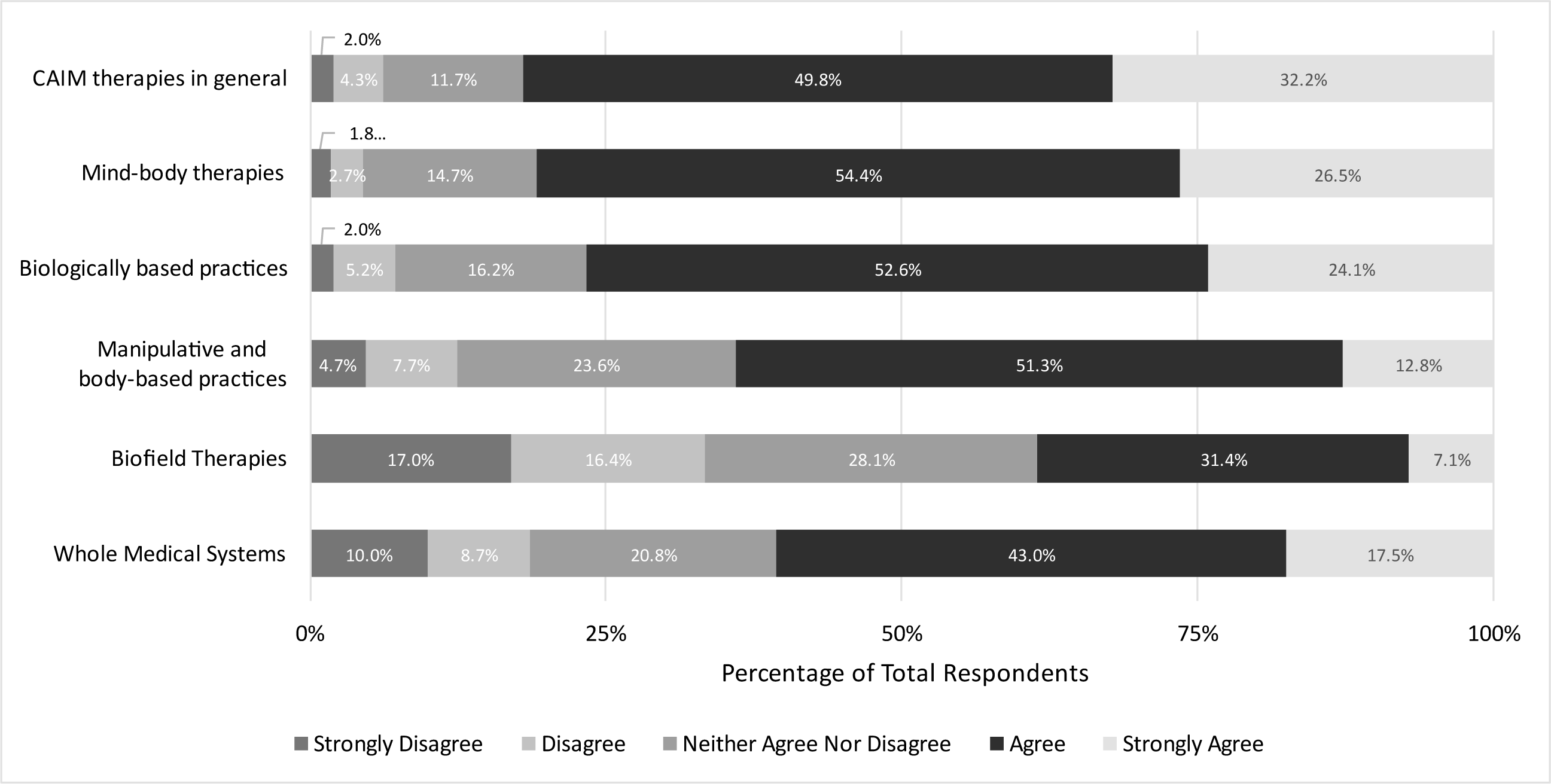
Participants’ Agreement Expressed Towards the Value of Conducting Research on Each CAIM Category.

### Thematic Analysis

When asked to share any remaining perceptions of CAIM at the end of the survey, a total of 28 codes were identified from the responses. The codes were then categorised under 7 major themes: CAIM research, CAIM categorisation, regulation and standardisation of CAIM, clinical practice of CAIM, CAIM’s challenges, CAIM’s benefits, and other. Some prevailing sub-themes were: more data on CAIM is required, as respondents strongly felt that there is currently a lack of evidence concerning CAIM’s safety and efficacy; broad categorisation of CAIM therapies is flawed, as respondents expressed that the grouping of CAIM treatments is overly general, and therefore, difficult to comment on; and finally, CAIM lacks standardisation and regulation, as respondents perceived CAIM to have poor quality-control surrounding its evidence, safety, and efficacy. Open-ended responses also revealed that participants believed CAIM research requires rigorous experimental design and analysis, and research funding should be selectively allocated towards promising CAIM therapies. Coding and thematic analysis data are available at: https://osf.io/s3nre.

## Discussion

The objective of this study was to explore neurology researchers and clinicians’ perceptions of CAIM to gather a comprehensive understanding of CAIM practices and awareness in clinical and research settings. The current literature on CAIM is predominantly based on patients’ perspectives; the majority of published literature that addresses clinicians’ standpoints largely encompasses other medical specialties^34^.

To the best of our knowledge, this study is the first to explore neurology researchers and clinicians’ perceptions of CAIM. Our findings demonstrate a considerable diversity of perceptions about the perceived safety, efficacy, and clinical promise of CAIM among neurology researchers and clinicians, both across and within individual CAIM categories. This heterogeneity may be attributed to a multitude of factors, including (but not limited to) age, gender, primary area of research, length of work experience, exposure to CAIM, and education on CAIM^34,37,39,51^. Multiple studies that investigate physicians’ attitudes towards CAIM for patients with epilepsy also suggest that cultural background has influence over physicians’ faith in the efficacy of CAIM therapies^52,53,54^. Our study found that overall, participants believed that CAIM is promising to treating neurological diseases and/or conditions. However, clinicians expressed that a lack of high-quality evidence is currently the most significant factor preventing CAIM from being implemented into routine clinical practices. This finding is in line with multiple studies that have also identified a global scepticism surrounding the safety and efficacy of CAIM due to a perceived lack of evidence and evidence-based resources in support of these therapies^37,39,42,55^. This perception is in line with previous literature that report a lower quantity of evidence regarding CAIM’s efficacy among patients with neurological disorders and/or conditions^10,16,19,34,39,40^. However, these studies also note that evidence is growing for a number of modalities, including mind-body therapy and biologically based practices^10^. Additionally, previous literature suggests that many physicians may not be aware of the few evidence-based resources that are currently available for CAIM^39^. While a lack of high-quality evidence does not necessarily imply that CAIM therapies are harmful, it is important to clarify any misconceptions about CAIM in order for physicians to comfortably advise patients about this category of therapies^37,54^. Furthermore, over 75% of respondents in this study agreed that there is value to conducting research on CAIM.

Our participants expressed concerns regarding the lack of standardisation and regulation of CAIM. Approximately 85% of respondents stated it is difficult to distinguish legitimate CAIM practices from scams or fraudulent claims. Currently, the evidence base for most CAIM modalities vary greatly in quality^56-59^. This is partly due to the fundamental differences between CAIM and conventional medicine that make it difficult to study many CAIM therapies under the gold-standard placebo-controlled randomised clinical trial^42,56,57^. The effects of certain CAIM therapies on neurological symptoms may also prove difficult to differentiate from non-specific treatment effects, for example, placebo and Hawthorne effects^56^. Although most respondents supported allocating more funding towards CAIM research, the open-ended data revealed that it is equally imperative to participants that careful selection and funding of studies takes place. Specifically, researchers and clinicians valued studies that employ rigorous methodology to ensure high-quality results. Previous literature indicates that this preference is expressed by other medical specialties as well^37,42,51,56^.

Among the five CAIM categories, our study found that neurology researchers and clinicians are the most open to mind-body therapies. Mind-body therapies consistently received the most positive sentiments across all CAIM categories regarding safety, efficacy, and clinical promise. Clinicians also felt the most comfortable recommending and counselling patients about mind-body therapies. Previous literature investigating CAIM practices among healthcare providers have reported similar findings. Multiple studies have reported that mind-body therapies—specifically, meditation and yoga—are commonly recommended by physicians^52,54,56,60^. These results may be attributed to the considerable evidence for mind-body modalities relative to other CAIMs, for example, biofield therapies^56^.

Our study revealed that most participating neurology clinicians have never received formal nor supplementary education about CAIM. This finding was unsurprising, as healthcare providers across all medical specialties consistently report that their lack of education on CAIM discourages them from recommending or counselling patients about this topic^37,39,40,51,60,61^. Similarly, in a study that investigated medical students’ attitudes towards CAIM, over 60% of students reported that the time devoted to CAIM in medical school felt inadequate. The study also found that students felt that their CAIM education was biased to be either pro- or anti-CAIM, as well as poor quality relative to the rest of their medical education^51^. This raises a concern surrounding patient-physician communication and patient-centric care. It is well-established in literature that the majority of CAIM users do not inform or consult their medical caregivers about their CAIM use^61-64^. Previous studies report that patients withhold CAIM-related information primarily due to a lack of inquiry by the provider as well as the perception that their provider is not knowledgeable about CAIM^64,65^. Parents of paediatric headache patients have admitted to being positively surprised by neurologists who acknowledge CAIM and are able to provide any insight towards integrating CAIM in their child’s treatment plan^65^. Given the increasing use of CAIM among patients with neurological conditions^6-28,34,38-42^, it is important that neurology clinicians inquire about CAIM use. To confidently coordinate safe, integrative treatment plans, it may be imperative that clinicians receive formal and/or supplementary education on CAIM^37,39,40,51,60,61^.

Although over half of our respondents agreed that clinicians should receive formal and supplementary education on CAIM, respondents’ opinions on whether CAIM should be integrated into mainstream medical practices were more heavily varied. This diversity is likely attributed to the key themes identified throughout our data, specifically: more high-quality evidence in support of CAIM is needed, CAIM lacks reliable standardisation and regulation, and CAIM can be a high-risk practice. These findings may be of value to medical educators and policy makers who can play an influential role in neurology researchers and clinicians’ awareness of CAIM^51,66^.

## Strengths and Limitations

Strengths of this study include the use of a cross-sectional survey due to its efficient and cost-effective nature^67^, which allowed us to gather data without requiring a long-term follow-up. This approach also allowed us to deliver the survey to an international sample and collect a greater range and diversity of perspectives about CAIM. A high completion rate was also achieved among those who responded. Limitations of this study include response bias, where researchers and/or clinicians who chose to participate in the survey may have different experiences with CAIM than those who opted out of the survey^67^. Additionally, the self-reporting nature of this study renders it susceptible to recall bias^67^. The low response rate and small sample size of the study may confound the representativeness of our results.

Furthermore, the data from this study was collected primarily from Europe and the Americas and may not be generalizable or representative of the global population of neurology researchers and/or clinicians. There was also a selection bias for English-speaking participants, further indicating that the actual representativeness of the study’s participants is uncertain. Finally, thematic analysis revealed that participants felt that the broad categorisation of CAIM therapies in our questionnaire was flawed, as respondents expressed that the grouping of different CAIM modalities was overly general and simplified, and therefore, difficult to comment on.

## Conclusions

This study investigated the practice and perceptions of CAIM among neurology researchers and clinicians. Acceptance of and experiences with CAIM varied across participants and within each individual CAIM category. Mind-body therapies were perceived to be the most promising to treating neurological disorders and was also the most accepted and practiced CAIM category among clinicians.

Despite growing patient demand, the current lack of scientific evidence for the safety and efficacy of CAIM discourages clinicians from implementing CAIM into routine clinical care. In addition, there is a general lack of knowledge about CAIM and CAIM research among neurology researchers and clinicians. The safety and efficacy of many CAIM therapies was highly ambiguous to participants. Previous literature suggests that clinicians’ scepticism may also be tied to the lack of CAIM discourse in formal and/or supplementary medical education. Our study provides a solid foundation to understanding researchers and clinicians’ perceptions of CAIM within the field of neurology. It establishes a compelling case for improving CAIM education and training efforts for medical professionals. Knowledge of how CAIM practices are perceived can help tailor educational resources and initiatives that better suit the needs of neurology researchers and clinicians, and in turn, encourages safe and informed patient care. Future work can build upon our study to monitor the implementation of CAIM education within the field of neurology.

## List of Abbreviations

CAIM: complementary, alternative, and integrative medicine
CHERRIES: Checklist for Reporting Results of Internet E-Surveys
MEDLINE: Medical Literature Analysis and Retrieval System Online
NLM: National Library of Medicine
NCCIH: National Center for Complementary and Integrative Health
OSF: Open Science Framework
PMID: PubMed identifier

## Declarations

## Ethics Approval and Consent to Participate

We sought and were granted ethics approval by the University Hospital Tübingen Research Ethics Board prior to beginning this project (REB Number: 325/2007 BO1).

## Consent for Publication

All authors consent to this manuscript’s publication.

## Availability of Data and Materials

All data and materials associated with this study have been posted on the Open Science Framework and can be found here: https://doi.org/10.17605/OSF.IO/HV3G5

## Competing Interests

The authors declare that they have no competing interests.

## Funding

This study was unfunded.

## Authors’ Contributions

JYN: designed and conceptualized the study, collected and analysed data, drafted the manuscript, and gave final approval of the version to be published.

SYL: assisted with the collection and analysis of data, made critical revisions to the manuscript, and gave final approval of the version to be published.

HC: assisted with the design and concept of the study and the analysis of data, made critical revisions to the manuscript, and gave final approval of the version to be published.

## Data Availability

All data and materials associated with this study have been posted on the Open Science Framework.

https://doi.org/10.17605/OSF.IO/HV3G5

## Notes

### Competing Interest Statement

The authors have declared no competing interest.

### Clinical Protocols

https://doi.org/10.17605/OSF.IO/R97MH

### Author Declarations

We sought and were granted ethics approval by the University Hospital Tubingen Research Ethics Board prior to beginning this project (REB Number: 325/2007 BO1).

